# Global Perspectives on Returning Genetic Research Results in Parkinson’s Disease

**DOI:** 10.1101/2024.07.06.24309029

**Authors:** Ai Huey Tan, Paula Saffie-Awad, Artur F Schumacher Schuh, Shen-Yang Lim, Harutyun Madoev, Azlina Ahmad-Annuar, Justin Solle, Claire Ellen Wegel, Maria Leila Doquenia, Sumit Dey, Maria Teresa Periñan, Mary B Makarious, Brian Fiske, Huw R Morris, Alastair J Noyce, Roy N. Alcalay, Kishore R. Kumar, Christine Klein, the Global Parkinson’s Genetics Program (GP2)

**Affiliations:** Division of Neurology, Department of Medicine, Faculty of Medicine, University of Malaya, Kuala Lumpur, Malaysia; CETRAM-Centro de Estudios de Trastornos del Movimiento, Clínica Santa María,Santiago, Chile; Departamento de Farmacologia, Universidade Federal do Rio Grande do Sul, Porto Alegre, Brazil; Serviço de Neurologia, Hospital de Clínicas de Porto Alegre, Porto Alegre, Brazil; Institute of Neurogenetics, University of Lübeck, Lübeck, Germany; Department of Biomedical Science, Faculty of Medicine, University of Malaya, Kuala Lumpur, Malaysia; The Michael J. Fox Foundation for Parkinson’s Research, New York, New York, USA; Department of Medical and Molecular Genetics, Indiana University, Indianapolis, Indiana, USA; Department of Neuroscience and Brain Health, Metropolitan Medical Center, Manila, Philippines; Centre for Preventive Neurology, Wolfson Institute of Population Health, Queen Mary University of London, London, UK; Unidad de Trastornos del Movimiento, Servicio de Neurología y Neurofisiología Clínica, Instituto de Biomedicina de Sevilla, Hospital Universitario Virgen del Rocío/CSIC/Universidad de Sevilla, Seville, Spain; Laboratory of Neurogenetics, National Institute on Aging, National Institutes of Health, Bethesda, MD, USA; Department of Clinical and Movement Neurosciences, UCL Queen Square Institute of Neurology, University College London, London, UK; Department of Neurology, Columbia University Irving Medical Center, New York, New York, USA; Movement Disorders Division, Neurological Institute, Tel Aviv Sourasky Medical Center, Tel Aviv, Israel and Tel Aviv School of Medicine, Tel Aviv University, Tel Aviv, Israel; Molecular Medicine Laboratory and Neurology Department, Concord Clinical School, Concord Repatriation General Hospital, The University of Sydney, Sydney, New South Wales, Australia; Translational Neurogenomics Group, Genomic and Inherited Disease Program, Garvan Institute of Medical Research, Darlinghurst, New South Wales, Australia; St Vincent’s Healthcare Campus, Faculty of Medicine, UNSW Sydney, Darlinghurst, New South Wales, Australia

## Abstract

In the era of precision medicine, genetic test results have become increasingly relevant in the care of patients with Parkinson’s disease (PD) and their families. While large PD research consortia are performing widespread genetic testing to accelerate discoveries, debate continues about whether, and to what extent, the results should be returned to patients. Ethically, it is imperative to keep participants informed, especially when findings are potentially actionable. However, research testing may not hold the same standards required from clinical diagnostic laboratories. The absence of universally recognized protocols complicates the establishment of appropriate guidelines. Aiming to develop recommendations on return of research results (RoR) practice within the Global Parkinson’s Genetics Program (GP2), we conducted a global survey to gain insight on GP2 members’ perceptions, practice, readiness, and needs surrounding RoR. GP2 members (*n*=191), representing 147 institutions and 60 countries across six continents, completed the survey. Access to clinical genetic testing services was significantly higher in high-income countries compared to low– and middle-income countries (96.6% vs. 58.4%), where funding was predominantly covered by patients themselves. While 92.7% of the respondents agreed that genetic research results should be returned, levels of agreement were higher for clinically relevant results relating to pathogenic or likely pathogenic variants in genes known to cause PD or other neurodegenerative diseases. Less than 10% offered separate clinically-accredited genetic testing before returning genetic research results. 48.7% reported having a specific statement on RoR policy in their ethics consent form, while 53.9% collected data on participants’ preferences on RoR prospectively. 24.1% had formal genetic counselling training. Notably, the comfort level in returning incidental genetic findings or returning results to unaffected individuals remains low. Given the differences in resources and training for RoR, as well as ethical and regulatory considerations, tailored approaches are required to ensure equitable access to RoR. Several identified strategies to enhance RoR practices include improving informed consent processes, increasing capacity for genetic counselling including providing counselling toolkits for common genetic variants, broadening access to sustainable clinically-accredited testing, building logistical infrastructure for RoR processes, and continuing public and healthcare education efforts on the important role of genetics in PD.

## INTRODUCTION

Over the past three decades, there has been accumulating evidence supporting an important role for genetics in the development and progression of Parkinson’s disease (PD),^1-3^ and genetic testing in PD is becoming more commonplace across clinical, research, and direct-to-consumer settings.

The Global Parkinson’s Genetics Program (GP2) is a major endeavour aiming to discover novel insights into the genetic drivers of PD, and to make this knowledge globally available and actionable. ^4^ This ambitious program aims to perform genotyping and/or sequencing in ∼200,000 individuals with PD and prioritizes the inclusion of populations worldwide that historically have been underrepresented in genetic studies.^5,6^ While the bulk of collected samples were initially planned to undergo genotyping using a single nucleotide polymorphism [SNP] array platform (i.e., the NeuroBooster Array),^7^ it is now anticipated that with the ongoing reduction in the costs associated with whole genome sequencing (WGS), many samples will be sequenced, thus increasing the power to detect nearly all forms of genomic variation in an unbiased manner.^8-10^

Currently, the yield of genetic testing in PD in most settings is ∼5-15%, depending on the population studied and the platform used (most commonly targeted gene panels or single-gene studies). ^11-13^ However, this has been shown to be as high as 40-50% in some populations. ^3,14-17^ Known PD/parkinsonism genes have either an autosomal dominant (e.g., *SNCA, LRRK2, VPS35*), autosomal recessive (e.g., *PRKN, PINK1, PARK7*/*DJ-1*) or X-linked (*TAF1*) mode of inheritance. Additionally, risk genes are recognized, and in particular carriers of *GBA1* variants have increased susceptibility to developing PD.^18^ New monogenic causes of PD continue to be discovered, such as *RAB32*, which was found in several populations in Africa, North America, and Europe.^19^ During the course of the GP2, it is expected that a large number of variants in PD genes with potential clinical relevance will be detected in a research setting.

The main purpose of a research program (such as the GP2) is to advance scientific understanding and gain mechanistic insights with the potential to benefit populations of people with PD.^20^ This is distinct from clinical testing, which is usually focused on attaining a diagnostic result which would then be used to inform clinical management.^20^ Traditionally, genetic results from research studies were not returned (for a variety of reasons discussed further below, including posing an “untenable burden on research infrastructure”, since disclosure can be resource-intensive),^20^ however, this practice is evolving. Practices for returning genetic research results also vary widely across different countries as a reflection of regional differences in the expertise and training of clinicians, the availability of genetic counselling resources, access to clinically accredited genetic testing, and attitudes of patients and the community.^21,22^ Some countries have adapted wide-scale research genetic testing such as the 100,000 Genomes Project in the United Kingdom, that later as the project developed, obtained diagnostic accreditation.^10^ Additionally, there are important ethico-legal considerations, which include the participant’s right of access to their personal data, the participant’s right to know and right not to know, and the researchers’ duty of care.^23^

The challenge of disclosing individual genetic findings to research participants presents both opportunities and risks, necessitating thoughtful consideration. Disclosing individual genetic research results can have direct benefits to participants, such as modifying medical management and providing more information regarding diagnosis and prognosis as well as opportunities for participation in clinical trials.^3,24,25^ Furthermore, there is a high level of interest and a general willingness of health professionals and researchers to return results, particularly if the results are thought to be clinically relevant and reliable.^8,24,26^ However, there are potential risks to returning research results, including the possibility of adverse psychological consequences to the participants and their family members – although some would argue that this has sometimes been overstated, and indeed positive implications on healthy behavior change have been reported.^27,28,29^ While GP2 strives for the highest quality of research results, the very stringent quality control measures that accredited diagnostic testing laboratories have to adhere to cannot usually be matched in the research setting, and errors such as mislabelling or mix-up of samples can occur.^30^ Moreover, despite the personal utility that individuals with PD derive from genetic test results, this area remains underexplored, especially in underrepresented and resource-constrained regions.^31^

Aiming to develop recommendations on return of results (RoR) practices within the GP2, we conducted a global survey to gain insights into the GP2 members’ perceptions, practices, readiness, and needs on returning results of genetic research testing. Here, we seek to better understand the demand for RoR and the potential challenges and risks of RoR in a diverse range of countries and settings. The results of the survey may help with the design of suitable approaches to return genetic research results to PD patients and families efficiently and safely, now and in the future.

## METHODS

### Development and execution of the GP2 Return of Results Survey

The survey was developed by six movement disorder neurologists with expertise in PD genetic testing from North and South America, Europe, Asia, and Australia, who are members of the GP2 RoR Interest Group (AHT, KK, PSA, AFSS, RA, and CK). The contents of the initial survey draft were discussed in online meetings. Each item of the survey was refined through two rounds of appraisal for content validity, relevance, clarity, and conciseness. The draft was then converted into an online format, accessible through different browsers and devices. Readability and usability of the online survey were tested by the working group members and an additional movement disorder neurologist and a medical geneticist. During each step, items that were unclear were revised accordingly. The online survey consisted of four sections of multiple-choice questions: A) Demographics, B) Access to genetic testing services in clinical practice, C) Perceptions and ethical considerations on returning genetic research results, and D) Readiness to return genetic research results (eAppendix).

Invitation to participate in the online survey was sent via email to 572 GP2 members including 415 GP2 investigators and 157 GP2 trainees (e.g., postgraduate students or trainees in related clinical, genetic and/or basic science GP2 projects) with two rounds of reminder emails. To improve the response rate, we addressed each GP2 member and explained the importance of the survey in developing a workflow for RoR in the GP2 in our invitation emails. Each GP2 member received an individualized survey link, which also enabled easy return to the survey at other times, until final submission. To ensure no missing survey data, each respondent was prompted to answer all the questions in one section before proceeding to the next section. A message of survey receipt and appreciation was sent upon submission of the survey. Descriptive data and chi-square analyses were conducted using the IBM SPSS ver.23.

## RESULTS

A total of 191 GP2 members representing 147 institutions and 60 countries across six continents completed the online survey between July 27, and August 17, 2023. The survey response rate was 39.3% (n=163/415) and 17.8% (n=28/157) in the GP2 investigator and trainee groups respectively. All submitted surveys had a 100% completion rate for each section. Respondent demographics are summarized in Figure 1. The highest numbers of respondents were from Asia (26.2%) and Europe (23.6%). Notably, 49.7% were from resource-limited regions (i.e., low– and middle-income countries (LMIC), as defined by the World Bank^22^). 71.3% were clinicians (101 movement disorder neurologists, 24 neurologists, and 11 other medical practitioners) while 22.5% were basic scientists/researchers and 5.2% were geneticists/genetic counsellors. More than two-thirds were working in university or academic teaching hospitals. Three-quarters (77.5%) of the respondents had >10 years of working experience in the healthcare field.

**Figure 1:**
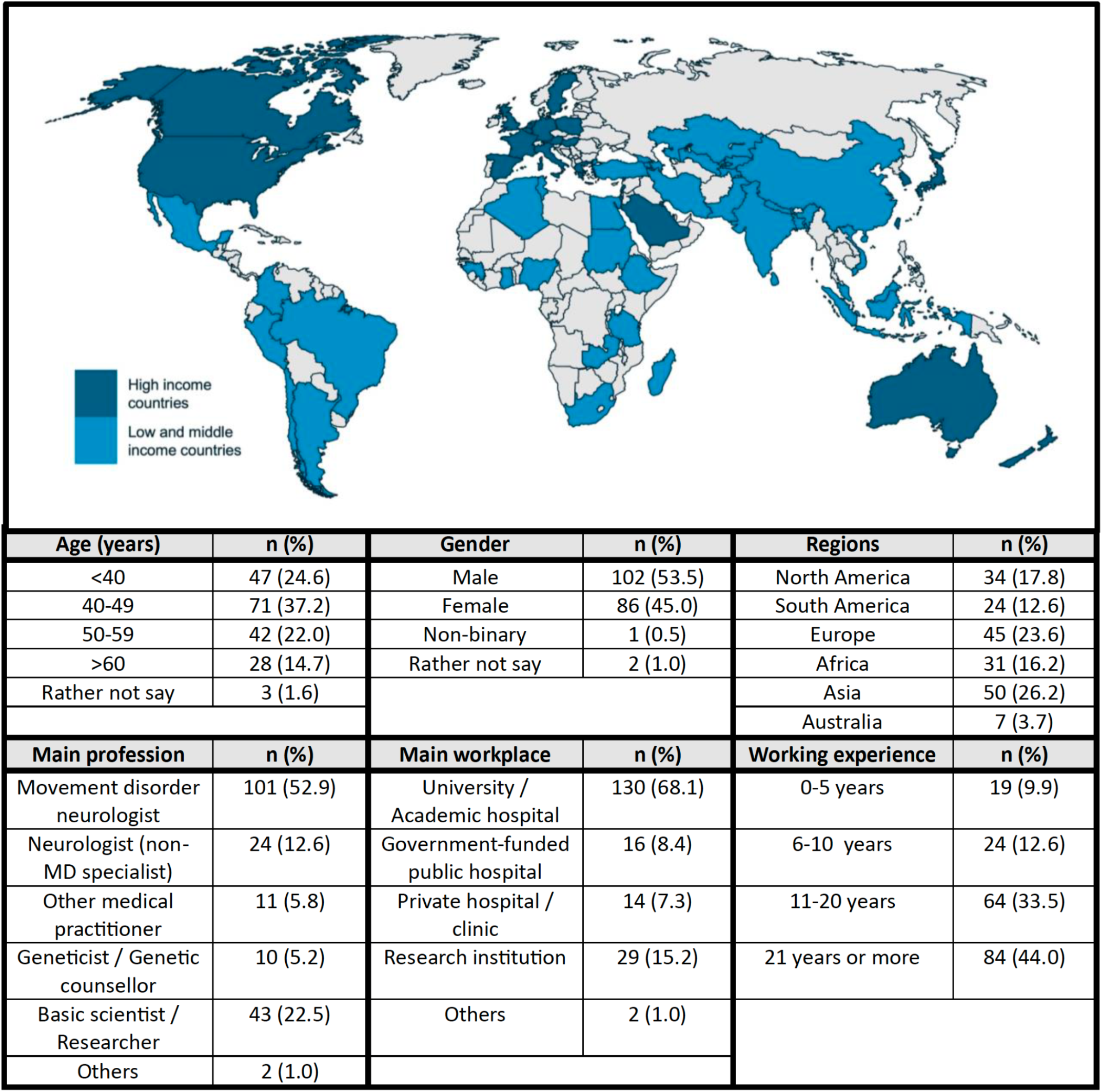
Demographics of 191 survey respondents. Highlighted in blue in the map are 60 countries represented by the survey respondents; high income countries are coloured in dark blue, while low and middle income countries are coloured in light blue. The table summarizes the age, gender, regions that the respondents originate from, main profession, main workplace and years of working experience in the healthcare field of the surveyed cohort. MD: Movement Disorder.

### Access to genetic testing in clinical practice

Among the 136 clinician respondents, 75% (n=102/136) reported having access to genetic testing in clinical practice, through clinical diagnostic laboratories in their institutions/countries (n=90/136) or outside their countries (n=34/136), or through genetic research laboratories (n=75/136). Table 1 depicts the differences in access to genetic testing and counselling services between respondents from high-income countries (HIC) vs. LMIC. Significantly larger proportions of respondents from HIC had access to genetic testing in clinical practice compared to those from LMIC (96.6% vs. 58.4%, p<0.001), where respondents from Africa and South America reported the lowest rates of access. Genetic testing in clinical practice was primarily paid for through government funding in HIC, while out-of-pocket payment was the primary funding mechanism for genetic testing in LMIC. Overall, 75.7% of clinician respondents (n=103/136) reported having access to genetic counselling services, with higher service availability in HIC vs. LMIC (88.1% vs. 66.2%, p=0.004). In LMIC, respondents reported higher access to genetic counselling services by neurologists or movement disorder neurologists (51.4%), compared to services by geneticists or genetic counsellors (35.7%). 25.4% of the respondents in HIC had access to genetic telemedicine services.

**Table 1:** Access to genetic testing and counselling services in clinical practice: Comparison between high income vs. low and middle income countries.

| Survey Items | Response from<br>all clinicians<br>(n=136) | Response from<br>clinicians in HIC<br>(n=59) | Response from<br>clinicians in<br>LMIC (n=77) | P value |
| --- | --- | --- | --- | --- |
|  | % answered yes | % answered yes | % answered yes |  |
| Access to genetic testing in clinical practice |  |  |  |  |
| Has access to genetic testing in clinical practice | 75.0 | 96.6 | 58.4 | <0.001* |
| Avenues for genetic testing in routine clinical practice |  |  |  |  |
| Clinical diagnostic lab in own institution | 38.2 | 67.8 | 15.6 | <0.001* |
| Research genetic lab in own institution | 41.2 | 71.2 | 18.2 | <0.001* |
| Other clinical diagnostic lab in own country | 52.9 | 69.5 | 40.3 | 0.001* |
| Other research genetic lab in own country | 22.8 | 33.9 | 14.3 | 0.008* |
| Clinical diagnostic lab outside the country | 25.0 | 28.8 | 22.1 | 0.426 |
| Research genetic lab outside the country | 21.3 | 25.4 | 18.2 | 0.399 |
| Funding for genetic testing in routine clinical practice |  |  |  |  |
| Government funding | 39.7 | 61.0 | 23.4 | <0.001* |
| Out-of-pocket funding (by patients) | 58.1 | 40.7 | 71.4 | <0.001* |
| Private insurance / prepaid funding | 33.8 | 42.2 | 27.3 | 0.071 |
| Development assistance funding | 4.4 | 3.4 | 5.2 | 0.697 |
| Access to genetic counselling services |  |  |  |  |
| Has access to genetic counselling services | 75.7 | 88.1 | 66.2 | 0.004* |
| Has access to genetic counselling by geneticist or genetic counsellor | 54.0 | 77.8 | 35.7 | <0.001* |
| Has access to genetic counselling by neurologist or MD neurologist | 58.1 | 66.7 | 51.4 | 0.101 |
| Has access to genetic telemedicine services | 14.0 | 25.4 | 5.2 | 0.001* |

HIC: High income countries; LMIC: Low and middle income countries. Comparisons between responses from HIC and LMIC were analysed using chi-square. \*Denotes statistical significance.

### Perceptions on and current practices in returning genetic research results

Figure 2 summarizes respondent perceptions on and current practices in returning genetic research results. 92.7% of the 191 respondents were of the opinion that individual genetic research results should be returned to research participants, although 52.9% felt that only clinically relevant results should be returned. 68.6% felt that genetic research results should be confirmed in a clinically-accredited diagnostic laboratory before being returned to participants, while 17.8% were unsure. A substantial majority (70.7-94.8%) felt that results regarding pathogenic or likely pathogenic variants in a gene known to cause PD or other neurodegenerative diseases, as well as variants known to increase the risk of PD (e.g., *GBA1* variants) should be returned. Slightly under half (47.1-48.2%) responded that ACMG-recommended incidental findings and negative results should be returned.

**Figure 2:**
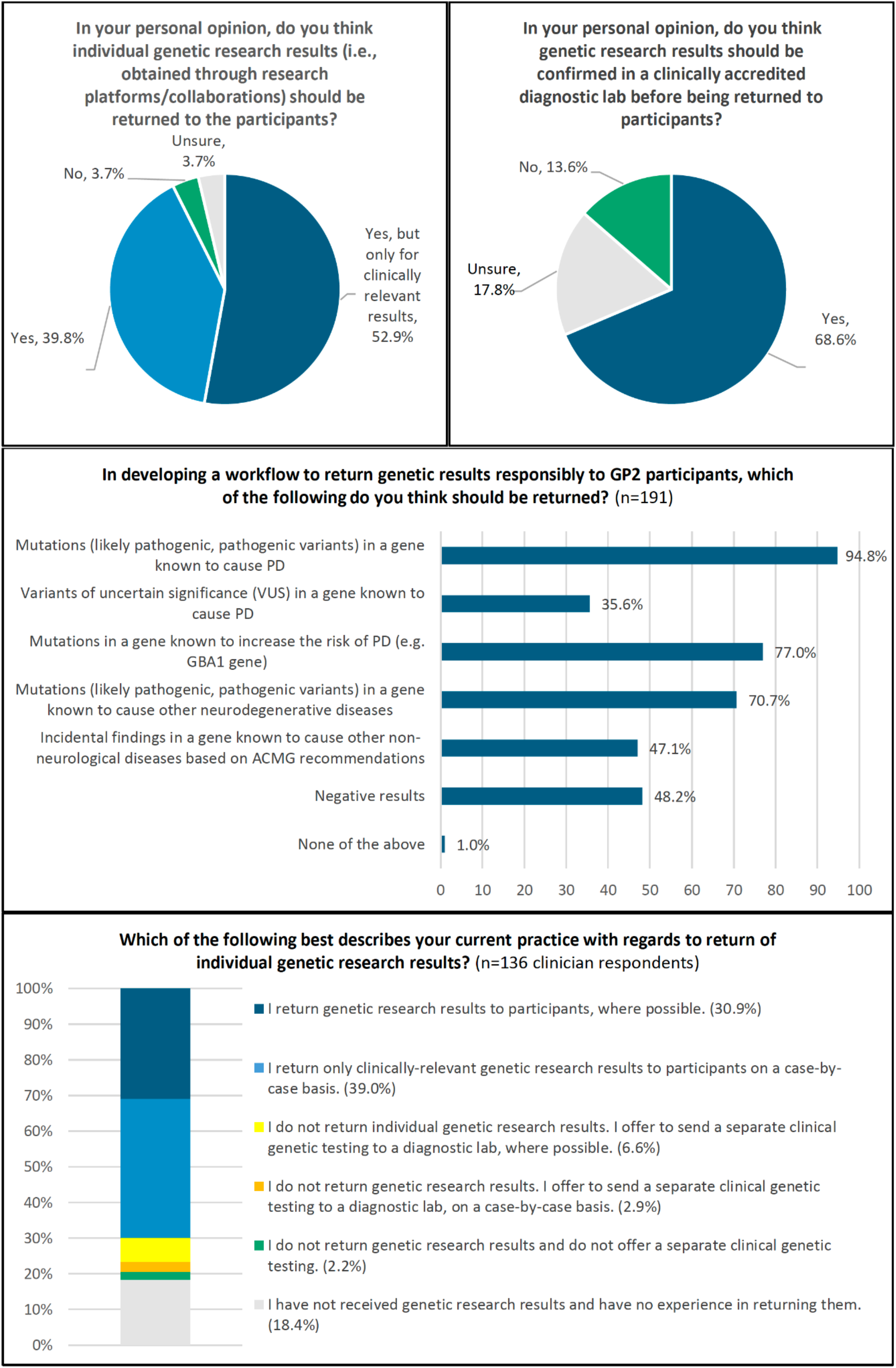
Perceptions and current practice on return of genetic research results

The majority of the clinician respondents (69.9%) practiced returning genetic research results directly to participants, while 9.5% offered separate clinical genetic testing through a diagnostic laboratory, and 2.2% did not return genetic research results or offer separate diagnostic testing. The top five major concerns in returning genetic research results included: 1) a lack of resources to validate genetic research results, 2) potential errors in genetic research results, 3) a lack of informed consent from research participants on RoR, 4) lack of pre-test genetic counselling during research recruitment, and 5) lack of experience/expertise in returning genetic results (Supplementary Table 1). With regard to potential implications on/issues surrounding research participants, the top five major concerns included: 1) possible impact on family members, 2) psychological consequences (e.g., stress, anxiety, and depression), 3) low health literacy and basic understanding of genetics, 4) lack of access to new therapeutics or clinical trials, and 5) potential negative impact on insurance (Supplementary Table 1).

### Ethics and local regulations on return of results

Out of 191 participants, 54 participants (28.2%) from 28 countries were aware of existing laws, policies, or guidelines governing or guiding RoR in their countries. Six participants from three countries stated that their local regulations do not allow RoR. 65 participants (34%) stated that there were no such local regulations, while the remaining 66 participants (34.6%) were unsure. There were instances of discordance between participants from the same country in their responses regarding the existence of local regulations, e.g., 9/29 participants from the USA considered that local regulations allowed RoR, four thought this was not permitted, six thought that there were no local regulations, and the remaining 10 were unsure.

A total of 93 participants from 73 institutions reported that their institutional ethics consent form contained a specific statement on RoR, whereby 37.0% could return genetic research results, 23.9% could return only clinically relevant results, 8.7% would obtain validation in a clinically-accredited laboratory before returning the results, 16.3% would not return research results, 4.3% had other RoR approaches, while 9.8% were unsure regarding their ethics statements on RoR. Although there was also some discordance in the responses by participants from the same institution regarding their institutional ethics statement, these were more consistent compared to the responses on local regulations. About half (53.9%) of the 191 respondents collected responses from participants during recruitment on whether they would like their genetic results to be returned, while 64.9% felt that the majority (>50%) of their participants would like to know their genetic results.

### Readiness to return genetic research results

Overall, 46 out of 191 respondents (24.1%) had formal training in genetic counselling, 31 of these respondents were clinicians. Among the 136 clinician respondents, the majority reported being comfortable in returning genetic research results (62.5% comfortable or very comfortable, 25% neutral, 9.6% slightly uncomfortable, 2.9% not comfortable). Notably, the proportion of clinician respondents who were comfortable or very comfortable returning results was higher among those who had formal training in genetic counselling (87.1% vs. 69.5%, p<0.001). Comfort levels in returning genetic research results differed according to different types of genetic variants (Figure 3). Overall, for affected individuals, most (>85%) respondents were comfortable returning clinically relevant pathogenic variants in PD genes, and >70% were comfortable returning the results on *GBA1* variants or pathogenic variants in other neurological disorder-related genes. Interestingly, the level of comfort in returning negative results was lower (45.8%-80%) than returning positive results in genes associated with PD or related neurological disorders (70.8%-100%). Only about 40% of movement disorder neurologists and non-MD neurologists were comfortable returning incidental findings to affected individuals. In general, a smaller proportion of respondents were comfortable returning results to unaffected individuals, about 70% were comfortable returning clinically relevant variants in PD genes, while about half were comfortable returning results on PD risk and causative variants in other neurologically-related genes.

**Figure 3:**
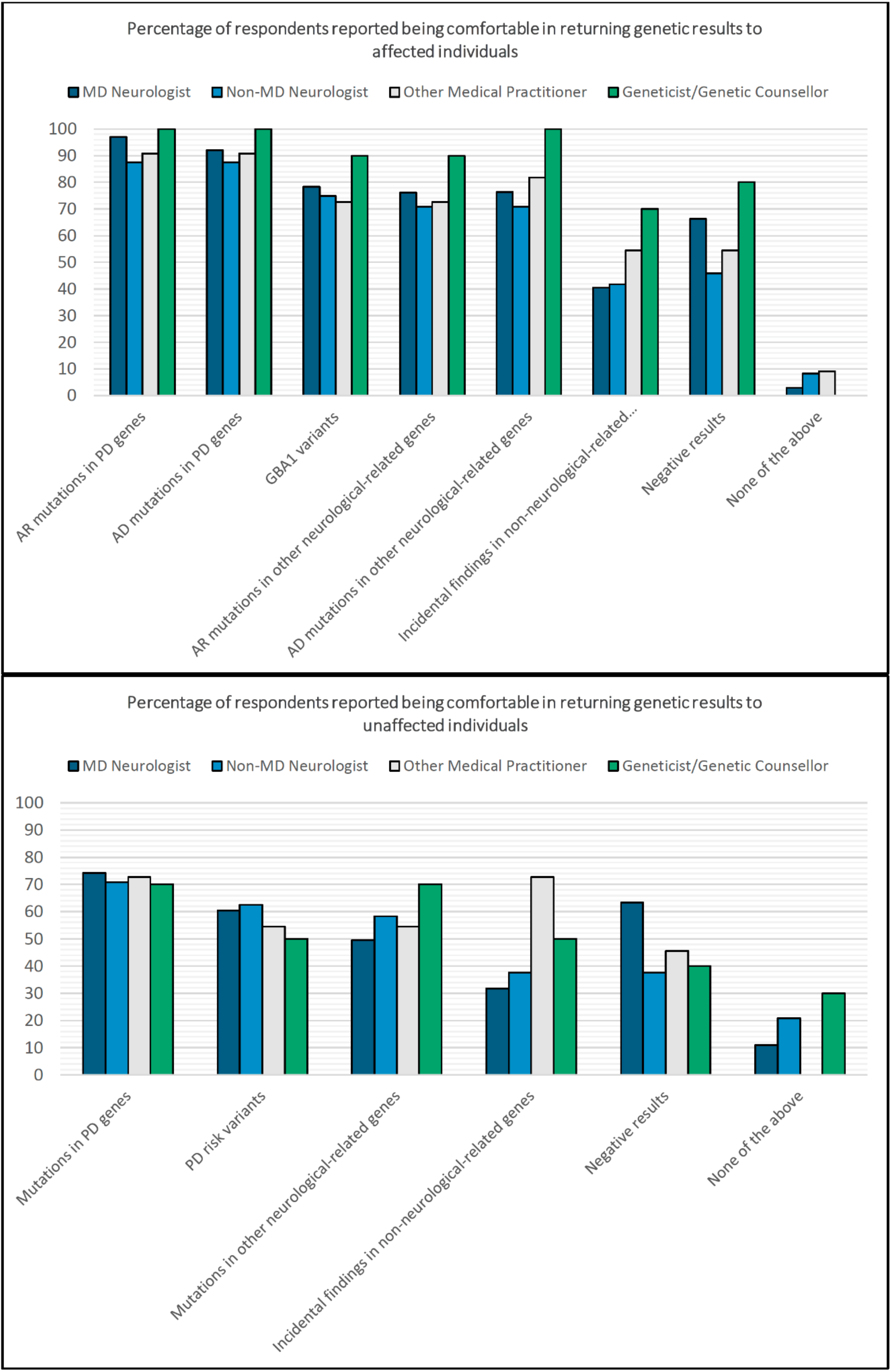
Comfort in returning genetic research results to affected and unaffected individuals

89% of the 136 clinician respondents reported the ability to recollect new samples from their research subjects for validation studies, and 49.6% estimated that they would be able to recollect samples from >50% of their cohort submitted to GP2. Among the respondents from HIC, 77.1% reported access to an accredited clinical diagnostic laboratory to validate selected GP2 results, while only 37.9% of respondents from LMIC reported similar access.

80.1% of the 191 respondents indicated a desire to receive additional information or training on how to return genetic research results. Among the different training platforms, certified training programs were the most preferred, followed by in-person training workshops, on-demand online modules, live online training courses, and digital reading materials.

## DISCUSSION

In this global survey of clinicians, researchers, and other professionals involved in PD genetics research, we uncovered novel insights and actionable findings in perceptions, practice, and readiness surrounding the return of genetic research results. Importantly, an overwhelming majority (>90%) of respondents felt that individual genetic research results should be returned, consistent with previous studies conducted among stakeholders and patients in genomics research. ^26,32^ The respondents divided on their view on the necessity of a clinical validation: two-thirds held the view that genetic research results should be validated in a clinically-accredited diagnostic lab, but only a very small proportion (<10% of the respondents) offered separate clinically-accredited genetic testing before RoR, likely reflecting current limitations in access to clinical genetic testing, and varying standards and practices around the world.^22^ We also identified important differences in access, resources, and training for genetic testing and validation, as well as ethical and regulatory considerations, between different institutions, countries, regions, and socio-economic strata. Formal training in genetic counselling is lacking and notably, the level of comfort in returning incidental genetic findings or returning results to unaffected individuals remains low.

52.9% of our survey respondents felt that only clinically relevant results (i.e., potentially diagnostic pathogenic/likely pathogenic variants in PD genes), rather than incidental or additional findings, should be returned. The potential impact on family members and psychological consequences were rated as top concerns. However, contrary to these common concerns, most participants from two large PD research cohorts in the USA reported no major adverse psychological effects from genetic result disclosure.^28,33^ In two separate studies, participants were prospectively offered choices regarding return of genomic results; 76.1-94.5% chose to learn all genetic results including incidental findings, while 5.5-14.4% chose a subset of results; only 0.5% of participants changed their choices after enrolment.^34,35^ While there may be hesitation in returning positive genetic results to unaffected individuals, in one survey, 46.1% of PD patients indicated they would have liked to know about their risk for PD, even in the absence of disease-modifying therapy.^36^ Taken together, strategies for RoR should embrace the heterogeneity of participants’ choices and personal preferences/values, as well as the evolving understanding on the impact of genetic results that may influence these choices (e.g., more defined knowledge on the natural history of the disease, penetrance, and treatment options). Dynamic forms of RoR consent allowing for changes in choices over time may be ideal,^34,37^ but will require more extensive allocation of resources to put into practice. Importantly, all involved should bear in mind that the ethical principle of autonomy also gives participants the right *not* to know their genetic result, and unwanted research information should never be forced onto participants.^8,24,26^

Ethical considerations in genomics research involve striking a balance between the potential benefits of returning individual genetic results (such as informing medical management or contributing to a participant’s understanding of their condition and assisting family and career planning, as well as enhancing research participation), and the risks (which include psychological harm, and the potential for misdiagnosis with lower quality control measures in research settings compared to clinical diagnostics). Legally, the situation is further complicated by country-specific requirements. For instance, in the USA, there are restrictions on the disclosure of results from laboratories that are not certified by the Clinical Laboratory Improvement Amendments (CLIA; through which the Centers for Medicare and Medicaid Services regulates human laboratory testing in the USA), highlighting the need for adherence to specific certification standards, which can be logistically impossible to be implemented within a global research program like the GP2. The legal framework varies significantly across different countries, reflecting disparities in expertise, the availability of genetic counselling resources, and access to clinically-accredited genetic testing. The lower access to genetic testing and related services in LMIC as highlighted in this survey and other previous reports^22,38,39^ may also influence local policies regarding RoR. For example, in some LMIC, the ethics committees may favour the disclosure of research results, even with their limitations, since this may be the only avenue available for testing to be done. In addition, there needs to be an awareness on the implications to other family members and future offspring including potential stigma that may be faced with genetic diagnosis in certain populations.^22^ Any approach to RoR must navigate this complex ethico-legal landscape, and support tailored strategies that respect local regulations and cultural sensitivities while striving to uphold the highest ethical and scientific standards in genetics research.

While there is now broader acceptance that there are many ethical and pragmatic reasons to return clinically-actionable genetic results, the practice of RoR raises several practical issues. While the majority of GP2 researchers expressed a willingness to return results, we found that only a quarter possessed formal genetic counselling training. Not surprisingly, a significant proportion found it challenging to return results on incidental and negative findings, or to unaffected individuals. Furthermore, about a quarter did not recognize the importance of confirming genetic research results in a clinically-accredited diagnostic laboratory prior to disclosure, and critically, access to such laboratories remains low in LMIC. There were also significant knowledge gaps among the respondents regarding their own local legal framework and ethical policy on RoR. Notably, only half ascertained their participants’ preferences on RoR during recruitment. These findings represent important gaps in RoR feasibility and readiness within the GP2 community. Based on these observations, we have formulated several recommendations for key next steps to improve RoR workflow in PD genetic research, starting from improvements to the informed consent process, to follow-up planning for RoR, summarized in Panel 1. While the GP2 is a genetic discovery initiative and not an effort primarily aiming to return genetic results to individual patients, we have also begun to navigate and support RoR by partnering with PDGENEration^40^, an initiative designed to carefully return genetic results to patients.

Online surveys offer many advantages including the opportunity to access a large sample of individuals worldwide, automation and consistency in the invitation language, cost and time efficiencies, and convenience for the respondents. By using a personalized email invitation and timely reminders, the response rate to this survey (almost 40%) was higher compared to others in the field (11-16%).^38^ Crucially, the survey cohort was representative of the professionals involved in PD genetics research, with participation from 147 institutions across 60 countries and six continents. Limitations of the survey include the sampling method (i.e., limited to only GP2 members), response bias (e.g., respondents may have more intention to return results compared to non-respondents), ambiguity when interpreting some questions, and limited depth (responses being based on multiple-choice format). Although this survey mainly targeted GP2 members, the ethical principles, perceptions and readiness for genetic testing and return of genetic research results are likely to be similar across various PD genetic research programs, therefore, increasing the generalizability of the results from this survey.

In conclusion, this survey highlights the diversity of perceptions, practice, resources, and readiness as well as ethical considerations surrounding the return of genetic research results, among professionals involved in PD genetics research worldwide. Recognizing these complexities and offering tailored strategies that address different needs and frameworks can pave the way for a more effective and ethically sound implementation of RoR, thereby advancing both genetics research and the delivery of personalized medicine.

## Data Availability

Data supporting the findings of this study are available from the corresponding author upon reasonable request.

## ACKNOWLEDGMENT

We thank our GP2 colleagues who contributed to the survey responses. We thank PDGENEration for their input and collaborations with regards to RoR policy and resources within the GP2.

## ETHICAL STATEMENT

This study received ethics approval from the University of Malaya Medical Centre Medical Research Ethics Committee (MREC ID NO: 2024625-13862).

## Panel 1: Suggested next steps to improve the ROR workflow in Parkinson’s genetic research

### Improved informed consent processes

This is particularly so for new centres with prospective cohort collection. Ethical documents should ideally have clear statements on ROR practices including the scope of findings to be returned, and should be compliant with local laws and regulations. Where possible, research participants should be given opportunities to indicate their preferences, including the option of choosing only certain types of findings (e.g., those considered “clinically relevant” to diagnosis, prognosis and family planning, and/or those considered “actionable” where prevention or treatment is available) to be returned, rather than the conventional “all or none” approach. It is also prudent to consider separate consent forms with clear and appropriate wordings for affected and unaffected research participants.

### Increased capacity for genetic counselling

This could include the creation of certified training programs, or less formalized in-person or online training courses. Regional centers for genetic counselling and creating networks for online counselling could also be considered. The development of genetic counselling toolkits for common genetic abnormalities in PD and related disorders could be helpful. Specific training resources should also be developed for counselling of unaffected individuals.

### Increased capacity to confirm results in a clinically-accredited laboratory

A cost-effective approach could involve regional collaboration to establish laboratories with local/regional certifications (or subsidized CLIA certification), thereby making the return of certified genetic results more feasible especially across lower-income regions. Research funding bodies for genetics research should consider funding the ROR processes, as these steps are also crucial in bolstering recruitment for genetics-informed clinical trials of disease-modifying therapies.

### Improved logistical infrastructure for “recontacting” and ROR processes

Researchers should be encouraged to develop a ROR plan as part of their research study design. Ideally, ethics approval should include provisions for the participants to be recontacted for repeat biological sampling for validation studies as well as participation in further related research (e.g., biomarker studies or clinical trials). Researchers should consider planning a clear pathway for the disclosure of validated genetic results (including who should do this, and when and how to return the results).

### Continuous efforts in educating healthcare professionals and the public about the role of genetics in Parkinson’s disease

This will foster a more informed and receptive environment for participation in genetics research and in the return of genetic research results.

## REFERENCES

1. Blauwendraat C, Nalls MA, Singleton AB. The genetic architecture of Parkinson’s disease. Lancet Neurol 2020;19:170–178.

2. Bandres-Ciga S, Diez-Fairen M, Kim JJ, Singleton AB. Genetics of Parkinson’s disease: An introspection of its journey towards precision medicine. Neurobiol Dis 2020;137:104782.

3. Lim SY, Klein C. Parkinson’s disease is predominantly a genetic disease. J Parkinson Dis 2024 Mar 25. doi: 10.3233/JPD-230376. Epub ahead of print.

4. Global Parkinson’s Genetics P. GP2: The Global Parkinson’s Genetics Program. Mov Disord 2021;36:842–851.

5. Lange LM, Avenali M, Ellis M, et al. Elucidating causative gene variants in hereditary Parkinson’s disease in the Global Parkinson’s Genetics Program (GP2). NPJ Parkinsons Dis 2023;9:100.

6. Towns C, Richer M, Jasaityte S, et al. Defining the causes of sporadic Parkinson’s disease in the global Parkinson’s genetics program (GP2). NPJ Parkinsons Dis 2023;9:131.

7. Bandres-Ciga S, Faghri F, Majounie E, et al. NeuroBooster Array: A Genome-Wide Genotyping Platform to Study Neurological Disorders Across Diverse Populations. medRxiv 2023.

8. Uffelmann E, Huang QQ, Munung NS, et al. Genome-wide association studies. Nature Reviews Methods Primers 2021;1.

9. McGuire AL, Gabriel S, Tishkoff SA, et al. The road ahead in genetics and genomics. Nat Rev Genet 2020;21:581–596.

10. Investigators GPP, Smedley D, Smith KR, et al. 100,000 Genomes Pilot on Rare-Disease Diagnosis in Health Care – Preliminary Report. N Engl J Med 2021;385:1868–1880.

11. Tysnes OB, Storstein A. Epidemiology of Parkinson’s disease. J Neural Transm (Vienna) 2017;124:901–905.

12. Trinh J, Lohmann K, Baumann H, et al. Utility and implications of exome sequencing in early-onset Parkinson’s disease. Mov Disord 2019;34:133–137.

13. Skrahina V, Gaber H, Vollstedt EJ, et al. The Rostock International Parkinson’s Disease (ROPAD) Study: Protocol and Initial Findings. Mov Disord 2021;36:1005–1010.

14. Lücking CB, Durr A, Bonifati V, et al. Association between early-onset Parkinson’s disease and mutations in the parkin gene. N Engl J Med 2000;342:1560–1567.

15. Lesage S, Durr A, Tazir M, et al. LRRK2 G2019S as a cause of Parkinson’s disease in North African Arabs. N Engl J Med 2006;354:422–423.

16. Rizig M, Bandres-Ciga S, Makarious MB, et al. Identification of genetic risk loci and causal insights associated with Parkinson’s disease in African and African admixed populations: a genome-wide association study. Lancet Neurol 2023;22:1015–1025.

17. Tay YW, Tan AH, Lim JL, et al. Genetic study of early-onset Parkinson’s disease in the Malaysian population. Parkinsonism Relat Disord 2023;111:105399.

18. Sidransky E, Nalls MA, Aasly JO, et al. Multicenter analysis of glucocerebrosidase mutations in Parkinson’s disease. N Engl J Med 2009;361:1651–1661.

19. Gustavsson EK, Follett J, Trinh J, et al. A pathogenic variant in RAB32 causes autosomal dominant Parkinson’s disease and activates LRRK2 kinase. medRxiv 2024.

20. Bredenoord AL, Kroes HY, Cuppen E, Parker M, van Delden JJ. Disclosure of individual genetic data to research participants: the debate reconsidered. Trends Genet 2011;27:41–47.

21. Saunders-Pullman R, Raymond D, Ortega RA, et al. International Genetic Testing and Counseling Practices for Parkinson’s Disease. Mov Disord 2023;38:1527–1535.

22. Tan AH, Cornejo-Olivas M, Okubadejo N, et al. Genetic Testing for Parkinson’s Disease and Movement Disorders in Less Privileged Areas: Barriers and Opportunities. Movement Disorders Clinical Practice 2023.

23. Vears DF, Hallowell N, Bentzen HB, et al. A practical checklist for return of results from genomic research in the European context. Eur J Hum Genet 2023;31:687–695.

24. Pont-Sunyer C, Bressman S, Raymond D, Glickman A, Tolosa E, Saunders-Pullman R. Disclosure of research results in genetic studies of Parkinson’s disease caused by LRRK2 mutations. Mov Disord 2015;30:904–908.

25. Keavney JL, Mathur S, Schroeder K, et al. Perspectives of people at-risk on Parkinson’s prevention research. J Parkinsons Dis. 2024 Mar 14. Epub ahead of print.

26. Vears DF, Minion JT, Roberts SJ, et al. Return of individual research results from genomic research: A systematic review of stakeholder perspectives. PLoS One 2021;16:e0258646.

27. Roberts JS. Assessing the Psychological Impact of Genetic Susceptibility Testing. Hastings Cent Rep 2019;49 Suppl 1:S38–S43.

28. Verbrugge J, Cook L, Miller M, et al. Outcomes of genetic test disclosure and genetic counseling in a large Parkinson’s disease research study. J Genet Couns 2021;30:755–765.

29. Frieser MJ, Wilson S, Vrieze S. Behavioral impact of return of genetic test results for complex disease: Systematic review and meta-analysis. Health Psychol. 2018 Dec;37(12):1134–1144.

30. Shevchenko Y, Bale S. Clinical Versus Research Sequencing. Cold Spring Harb Perspect Med 2016;6.

31. Pal G, Cook L, Schulze J, et al. Genetic Testing in Parkinson’s Disease. Mov Disord 2023;38:1384–1396.

32. Hill EJ, Robak LA, Al-Ouran R, et al. Genome sequencing in the Parkinson disease clinic. Neurol Genet. 2022;8(4):e200002.

33. Cook L, Verbrugge J, Schwantes-An TH, et al. Providing genetic testing and genetic counseling for Parkinson’s disease to the community. Genet Med 2023;25:100907.

34. Hoell C, Wynn J, Rasmussen LV, et al. Participant choices for return of genomic results in the eMERGE Network. Genet Med 2020;22:1821–1829.

35. Bishop CL, Strong KA, Dimmock DP. Choices of incidental findings of individuals undergoing genome wide sequencing, a single center’s experience. Clin Genet 2017;91:137–140.

36. Schaeffer E, Rogge A, Nieding K, et al. Patients’ views on the ethical challenges of early Parkinson disease detection. Neurology 2020;94:e2037–e2044.

37. Mackley MP, Fletcher B, Parker M, Watkins H, Ormondroyd E. Stakeholder views on secondary findings in whole-genome and whole-exome sequencing: a systematic review of quantitative and qualitative studies. Genet Med 2017;19:283–293.

38. Gatto EM, Walker RH, Gonzalez C, et al. Worldwide barriers to genetic testing for movement disorders. Eur J Neurol 2021;28:1901–1909.

39. Schumacher-Schuh AF, Bieger A, Okunoye O, et al. Underrepresented Populations in Parkinson’s Genetics Research: Current Landscape and Future Directions. Mov Disord 2022.

40. Parkinson’s Foundation. PD Generation: Mapping the Future of Parkinson’s disease. Available from https://www.parkinson.org/advancing-research/our-research/pdgeneration. Accessed 22 April 2024.

