## Supplementary material for "Global Perspectives on Returning Genetic Research Results in Parkinson’s Disease": Supp table 1

**Supplementary Table 1: Concerns regarding return of genetic research results (n=191)**

| | Percentage of respondents' rating (%) | | |
|---|---|---|---|
| **Concerns** | **No concern** | **Minor concern** | **Major concern** |
| **General concerns on returning genetic research results** | | | |
| Lack of pre-test genetic counselling during research recruitment | 19.4 | 43.5 | **37.2** |
| Lack of informed consent from research participants on return of research results | 25.7 | 36.6 | **37.7** |
| Potential error in genetic research results | 10.5 | 46.1 | **43.5** |
| Lack of resources to validate genetic research results | 14.7 | 40.8 | **44.5** |
| Lack of experiences/expertise in returning genetic results | 26.7 | 36.6 | **36.6** |
| Lack of time in returning genetic results | 37.7 | 38.2 | 24.1 |
| Potential negative implications to research participants | 19.9 | 45.5 | 34.6 |
| Ethics approval does not permit return of genetic research results | 44.5 | 24.6 | 30.9 |
| Governmental/Medical regulations do not permit return of genetic research results | 52.9 | 23.0 | 24.1 |
| **Concerns regarding potential implications on research participants** | | | |
| Low health literacy and basic understanding of genetic results | 8.4 | 41.4 | **50.3** |
| Psychological impact (e.g., stress, anxiety, depression) | 4.2 | 36.6 | **59.2** |
| Socio-cultural impact (e.g., social stigma) | 13.6 | 55.5 | 30.9 |
| Potential negative impact on employment | 26.7 | 48.2 | 25.1 |
| Potential negative impact on insurance | 29.3 | 38.2 | **32.5** |
| Potential Implications to other family members | 4.7 | 33.0 | **62.3** |
| Low clinical utility of genetic results in changing management | 21.5 | 48.7 | 29.8 |
| Lack of access to new therapeutics or clinical trials | 17.3 | 36.6 | **46.1** |
| Lack of genetic data privacy and confidentiality policy | 33.5 | 36.6 | 29.8 |

Highlighted in bold are the top five major concerns in each category.
