## Supplementary material for "Global Perspectives on Returning Genetic Research Results in Parkinson’s Disease": Supp file

Supplementary File 1

### RETURN OF GENETIC RESEARCH RESULTS IN GP2

This 10-minute survey is an initiative led by GP2, designed to gain insights on how our community addresses returning results of genetic research testing. Your valuable input will help us identify your needs and requirements to develop an efficient workflow to return genetic research results in GP2.

#### Section A. Demographics

1. Age:
- Less than 30 years old
- 30 to 39 years old
- 40 to 49 years old
- 50 to 59 years old
- 60 years or older
- Rather not say

2. Gender:
- Female
- Male
- Non-binary/diverse
- Rather not say

3. Which country are you based in? (List of countries as a drop-down list)

4. Which of the following best describes your profession? (One option only)
- Movement disorders neurologist
- Neurologist (non-movement disorders specialist)
- Medical geneticist/clinical geneticist
- Basic scientist
- Medical doctor
- Genetic counsellor
- Research nurse
- Others, please specify: (free text box)

5. How long have you been working as a healthcare professional or in a healthcare field?
- 0 to 5 years
- 6 to 10 years
- 11 to 20 years
- 21 years or more

6. Which of the following best describes your workplace? (One option only)
- University/Academic Teaching Hospital
- Government-Funded Public Hospital
- Private Hospital/Clinic
- Research Institution/Centre
- Others, please specify: (free text box)

#### Section B. Access to genetic testing services in clinical practice

7.1. Do you have access to genetic testing in your clinical practice? (One option only)

- Yes
- No

7.2. If yes, please describe your genetic testing access platforms (please tick all that apply).
(This question will only appear if answer Yes to 7.1. Multiple options allowed.)
- Clinical diagnostic genetic labs in my institution
- Research genetic labs in my institution
- Other clinical diagnostic genetic labs/companies in my country
- Other research genetic labs in my country
- Other clinical diagnostic genetic labs/companies outside my country
- Other research genetic labs outside my country

8. Outside the research context, how are genetic tests usually funded in your clinical practice? (please tick all that apply) (Multiple options allowed)
- Government funding
- Out-of-pocket funding
- Prepaid private funding
- Private insurance
- Development assistance funding
- Others; please specify: (free text box)

9.1. Do your patients have access to genetic counselling (e.g., by a geneticist/genetic counsellor, neurologist or other trained healthcare professional)?
- Yes
- No

9.2. If yes, please share with us information regarding the nature of the genetic counselling service in your institution (please tick all that apply).
(This question will only appear if answer Yes to 9.1. Multiple options allowed.)
- Genetic counselling is provided by a medical geneticist/genetic counsellor
- Genetic counselling is provided by a neurologist/movement disorder specialist
- Genetic counselling is provided by a certified and trained healthcare professional, but, not a medical geneticist/genetic counsellor or neurologist/movement disorder specialist
- Genetic counselling is provided as in-person consultation in my institution.
- Genetic counselling is provided via telephone or video telemedicine service.
- Patients are referred outside my institution to receive genetic counselling
- Genetic counselling is provided prior to the genetic test (pre-test)
- Genetic counselling is provided after obtaining the genetic test results (post-test)

#### Section C. Perceptions and ethical considerations on returning genetic research results

10. In your personal opinion, do you think individual genetic research results (i.e., obtained through research platforms/collaborations) should be returned to the participants? (One option only)
- Yes
- Yes, but only for clinically relevant genetic results
- No
- Unsure

11. In your personal opinion, do you think genetic research results should be confirmed in a clinically-accredited diagnostic lab before being returned to participants? (One option only)

- Yes
- No
- Unsure

12. Which of the following best describes your current practice with regards to return of individual genetic research results? (One option only)

- I return genetic research results to participants, where possible.
- I return only clinically relevant genetic research results to participants on a case-by-case basis
- I do not return individual genetic research results. I offer to send a separate clinical genetic testing to a diagnostic lab, where possible.
- I do not return genetic research results. I offer to send a separate clinical genetic testing to a diagnostic lab, on a case-by-case basis.
- I do not return genetic research results and do not offer a separate clinical genetic testing.
- I have not received genetic research results and have no experience in returning them.

13. What are your main concerns regarding return of genetic research results? Please rate the statements below. (For each point below, select one of 3 options: No Concern, Minor Concern, Major Concern)

- Lack of pre-test genetic counselling during research recruitment
- Lack of informed consent from research participants on return of research results
- Potential error in genetic research results
- Lack of resources to validate genetic research results
- Lack of experiences/expertise in returning genetic results
- Lack of time in returning genetic results
- Potential negative implications to research participants
- Ethics approval does not permit return of genetic research results
- Governmental/Medical regulations do not permit return of genetic research results

14. With regards to potential implications of genetic results on research participants, what are your main concerns? Please rate the statements below. (For each point below, select one of 3 options: No Concern, Minor Concern, Major Concern)

- Low health literacy and basic understanding of genetic results
- Psychological impact (e.g., stress, anxiety, depression)
- Socio-cultural impact (e.g., social stigma)
- Potential negative impact on employment
- Potential negative impact on insurance
- Potential Implications to other family members
- Low clinical utility of genetic results in changing management
- Lack of access to new therapeutics or clinical trials
- Lack of genetic data privacy and confidentiality policy

15. Does your country have laws/policies/guidelines regarding the return of genetic research results? (One option only)

- Yes, and they indicate that genetic research results can be returned to participants
- Yes, and they indicate that genetic research results cannot be returned to participants
- No, there is no law/policy/guideline regarding the return of genetic research results in my country
- Unsure
- Others, please specify: (free text box)

16.1. Is there a specific statement on the consent form approved in your institution on whether genetic research results can be returned to GP2 participants? (One option only)

- ○ Yes
- ○ No

16.2. If yes, what does the statement in the consent form say?
(This question will only appear if answer Yes to 15.1. One option only)

- ○ Genetic research results can be returned to the participants
- ○ Only clinically relevant genetic research results can be returned to the participants
- ○ Clinically relevant genetic research results will be validated in an accredited lab and returned to the participants
- ○ Genetic research results will not be returned to the participants, however options for clinical genetic testing will be offered based on the genetic research results.
- ○ Genetic research results will not be returned to the participants
- ○ Unsure
- ○ Others, please specify (you can copy the statement from your consent form): (free text box)

17. In your recruitment efforts for GP2, were participants asked if they would like their genetic results to be returned? (One option only)

- ○ Yes
- ○ No
- ○ Unsure

18. In your opinion, what proportion of your GP2 participants would like to receive back their genetic results? (One option only)

- ○ <25%
- ○ 25-50%
- ○ 50-75%
- ○ >75%
- ○ Unsure

19. In developing a workflow to return genetic results responsibly to GP2 participants, which of the following do you think should be returned? (please tick all that apply) (Multiple options allowed)

- ○ Mutations (likely pathogenic, pathogenic variants) in a gene known to cause PD
- ○ Variants of uncertain significance (VUS) in a gene known to cause PD
- ○ Mutations in a gene known to increase the risk of PD (e.g. *GBA* gene)
- ○ Mutations (likely pathogenic, pathogenic variants) in a gene known to cause other neurodegenerative diseases
- ○ Incidental findings in a gene known to cause other non-neurological diseases, according to the ACMG recommendations
- ○ Negative results
- ○ None of the above
- ○ Others, please specify: (Provide free text box)

#### Section D. Readiness to return genetic research results

20. Have you had any formal training in counselling your patients on genetic findings? (One option only)

- ○ Yes

- No

21. Do you feel comfortable returning results from GP2? (One option only)
- Very comfortable
- Comfortable
- Neutral
- Slightly uncomfortable
- Not comfortable.

22. What type of results would you be comfortable returning from the GP2?

22.1 For affected individuals (Please tick all that apply)
- Clinically relevant variants in known autosomal recessive PD genes (e.g., PRKN, PINK1, DJ1)
- Clinically relevant variants in known autosomal dominant PD genes (e.g., LRRK2, VPS35, SNCA)
- GBA variants
- Clinically relevant variants in other autosomal recessive genes related to the neurological condition (e.g., SPG11, SPG7)
- Clinically relevant variants in other autosomal dominant genes related to the neurological condition [e.g., ATXN2 (SCA2), HTT (Huntington's disease), C9orf72, MAPT, GCH1]
- Clinically relevant incidental findings in genes not related to the neurological condition
- Negative results
- None

22.2 For unaffected individuals (Please tick all that apply)
- Clinically relevant variants in known PD genes
- PD genetic risk variants
- Clinically relevant variants in other genes related to a neurological condition
- Clinically relevant incidental findings in genes not related to a neurological condition
- Negative results
- None

23. With regards to validating genetic research results from GP2, would you have access to an accredited clinical diagnostic lab in your country to validate selected variant/gene(s) from the GP2 study? (One option only)
- Yes
- No
- Unsure

24.1. With regards to validating genetic research results, would you be able to collect new samples from a proportion of your GP2 research participants? (One option only)
- Yes
- No

24.2. If yes, please indicate what proportion of your GP2 participants would you be able to resample? (This question will only appear if answer Yes to 24.1. One option only)
- <10%
- 10-25%
- 25-50%
- 50-75%
- >75%

25.1. Would you like to receive further information and training on how to return results for the GP2 study? (One option only)

- Yes
- No

25.2. If yes, what form of training platform will you find most helpful? (Please rate the following items: 1: Not helpful, 2: Somewhat helpful, 3: Helpful; 4: Very Helpful) (This question will only appear if answer Yes to 25.1)

- In-person training workshop
- Live virtual training course
- On-demand online modules
- Digital reading materials
- Certified training program

Thank you very much for your kind contributions towards this important effort in GP2.

Many thanks and our very best wishes,
GP2 Return-Of-Results Working Group
